# Association between lifestyle at different life periods and brain integrity in older adults

**DOI:** 10.1101/2024.01.31.24302049

**Authors:** Anne-Laure Turpin, Francesca Felisatti, Florence Mézenge, Brigitte Landeau, Denis Vivien, Vincent de la Sayette, Gaël Chételat, Julie Gonneaud, the Medit-Ageing Research Group

## Abstract

**Importance:** Lifestyle factors have been associated with dementia risk and neuroimaging markers of ageing and Alzheimer’s Disease (AD), but the period at which they have the greatest influence remains unclear.

**Objective:** To determine the relative influence of lifestyle at different life periods on older adults’ brain health.

**Design, Setting, and Participants:** Baseline data from the Age-Well trial were used in this study. Cognitively unimpaired participants aged 65 years and older were recruited in the general population between November 2016 to March 2018 in Caen, France. Analysis took place between June 2022 and September 2023.

**Exposure(s):** The Lifetime of Experiences Questionnaire (LEQ) was used to assess lifestyle during young adulthood (3-30y), midlife (30-65y) and late-life (>65y). For each life period, LEQ score is divided into specific and non-specific subscores.

**Main Outcome(s) and Measure(s):** Multiple regressions were conducted including lifestyle at the three life periods (in the same model) to predict gray matter volume (GMv; from structural MRI), glucose metabolism (FDG-PET), perfusion (Florbetapir-PET, early-acquisition) and amyloid burden (Florbetapir-PET, late-acquisition), both in AD-sensitive regions and voxel-wise, controlling for age and sex. Then, comparisons of correlations between lifestyle at each life period, as well as between specific versus non-specific activities, and neuroimaging outcomes were performed.

**Results:** Of the 135 older adults (mean age=69.3±3.79), 61.5% were women. No significant association was found between LEQ scores and AD-sensitive regions. While LEQ-young was not associated with neuroimaging, LEQ-midlife was more strongly associated with GMv, including in the anterior cingulate cortex, and with amyloid burden in the precuneus compared to the other periods. LEQ-late showed stronger associations with perfusion and glucose metabolism than LEQ-young and LEQ-midlife in medial frontal regions. Lower amyloid burden was more strongly correlated with LEQ-midlife specific than non-specific activities (z=-2.0977, p<.05, [95% CI, −0.3985 - −0.0102]) while perfusion was more strongly correlated with LEQ-late non-specific than specific scores (z=2.4369, p<.01, [95% CI, 0.0.415-0.4165]).

**Conclusions and Relevance:** Lifestyle at different life periods might have complementary benefits on structural/molecular *versus* functional markers of brain health in late-life. Interestingly, these associations were found in regions related to reserve/resilience and aging.

**Trial registration:** Clinicaltrials.gov Identifier: NCT02977819

## Introduction

Ageing is a physiological and complex process occurring throughout life that affects the body and organs, including the brain, and increases vulnerability to various diseases, including a higher risk of dementia. The most common form of dementia is Alzheimer’s Disease (AD) (Breijyeh & Karaman, 2020; Crous-Bou et al., 2017) biologically characterized by the presence of amyloid plaques, neurofibrillary tangles, as well as neuronal and synaptic loss. One of the major challenges consists to prevent the disease and associated brain alterations.

There’s a large heterogeneity in aging that could be explained by differences in resilience (Stern et al., 2020, 2023), including differences in i) cognitive reserve, the brain’s ability to cope with cerebral damage using cognitive processes and brain networks, ii) brain reserve, reflecting the neurobiological capital differing among people allowing some of them to better deal with brain ageing and pathology before symptoms onset or iii) brain maintenance, related to the absence of biological brain changes allowing a preservation of cognition in old age. These mechanisms could be influenced by a variety of genetic and environmental factors, including lifestyle, throughout life. In that context, lifestyle could be a promising target to improved cognitive and brain health across life and prevent, or at least delay, dementia.

Consistently, a healthier lifestyle is commonly associated with longer life expectancy (Chudasama et al., 2020; Li et al., 2018) and greater brain and cognitive outcomes in cognitively unimpaired older adults (Bradley et al., 2022; Heger et al., 2021; Melzer et al., 2021). The latest report of the Lancet Commission (Livingston et al., 2020), suggesting that ∼40% of dementia cases could be prevented by acting on various modifiable risk factors, proposes a certain temporality of action of these different factors (e.g. education during early-life, cardiovascular factors at midlife and physical inactivity or social isolation in late-life). This raises the question of the life periods at which lifestyle could have its greatest benefit on brain health, which would be critical to tailor effective lifestyle-based prevention strategies. Several studies have already assessed the associations between lifestyle factors at different life periods, assessed separately, and brain outcomes. Such assessment doesn’t allow to consider the shared variance of lifestyle across the lifespan, nor to directly compare the effect of life periods between them. Additionally, only one neuroimaging modality is usually investigated, giving a fragmented view of the associations between lifestyle through lifespan and brain integrity and their potential mechanisms (Chen et al., 2019; Kim et al., 2018; Landau et al., 2012).

The present study aims at further understanding the associations between lifestyle at different life periods and brain markers of aging and AD in cognitively unimpaired older individuals. More specifically, we aim at identifying the relative associations of lifestyle engagement in early, mid and late adulthood with brain outcomes, using multimodal neuroimaging in adults aged 65 and over. We hypothesized that lifestyle behaviors do not have the same influence on older adults’ brain health depending on the life period, which will result in association with brain outcomes that will differ between the different life periods.

## Materials & methods

### Participants

The Age-Well randomized clinical trial included 137 cognitively unimpaired older adults aged over 65 years at baseline (Poisnel et al., 2018). Individuals were recruited from the general population from November 2016 to March 2018 in Caen, France (see Flow Chart - Supplementary Figure 1). Inclusion and exclusion criteria are fully detailed elsewhere (Poisnel et al., 2018). Briefly, all participants were native French speakers, retired for at least 1 year, had at least 7 years of education, and performed within the normal range on standardized cognitive tests. Two participants were excluded for not meeting major eligibility criteria (Flow Chart, Supplementary Figure 1 for details). As a result, 135 participants were included and underwent lifestyle assessment along with multimodal neuroimaging at baseline (Figure 1A). Participants’ characteristics are presented in Table 1.

**Figure 1:**
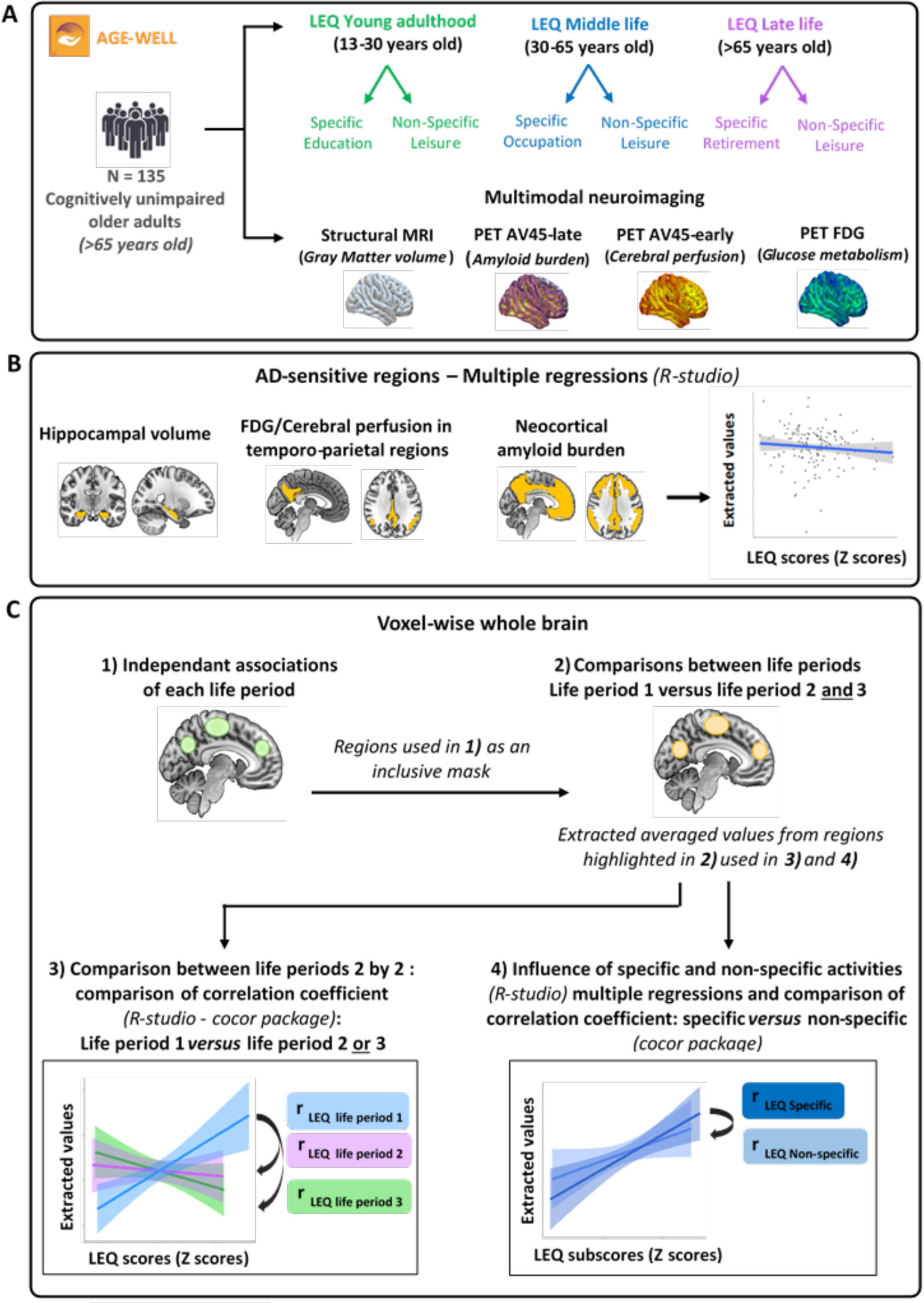
Design overview. A. 135 cognitively unimpaired older adults from the Age Well cohort completed the Lifetime of Experiences Questionnaire (LEQ) at baseline, allowing a retrospective evaluation of their lifestyle during young adulthood, middle life and late-life. A score was obtained for each life period and reflected both activities that were specific and non-specific to this period. Multimodal neuroimaging data were also obtained at baseline, including structural MRI (gray matter volume), FDG-PET (cerebral glucose metabolism) and Florbetapir-PET scans (early frames/cerebral perfusion and late frames/amyloid burden). B. Multiple linear regressions were performed including LEQ scores for the three life periods as predictors of each AD-signature regions (one model per imaging modality; hippocampal volume of the, temporo-parietal glucose metabolism and cerebral perfusion, neocortical amyloid burden); controlling for age and sex. C. Whole-brain analyses: 1) voxel-wise multiple linear regressions were conducted to assess the independent associations of LEQ score at each life period (in the same model) with each neuroimaging modalities (in separate models). Results were considered significant at p<.005 (uncorrected). When an association was found between lifestyle at a given life period and imaging in a set of regions, we conducted additional analyses to further understand the specificity of this association and the type of activity that could drive this effect. 2) Within the regions previously evidenced (Figure 1C1, green regions) we assessed whether the association with this life period was stronger than the association with the 2 other life periods. Volume/SUVR of the regions found to be more related to this life period than to the 2 others (Figure 1C2, yellow regions) were extracted to 3) conduct a 2-by-2 correlation coefficient comparisons between the three life periods and 4) assess the influence of specific versus non-specific activities conducting comparison of the correlation coefficient between specific and non-specific subscores. Abbreviations: LEQ: Lifetime of Experiences Questionnaire; MRI: Magnetic Resonance Imaging; PET: Positron Emission Tomography; SUVR: Standardized Uptake Value Ratio

The Age-Well trial was sponsored by Inserm and received approval from regional ethics committee (Comité de Protection des Personnes Nord-Ouest III, Caen, France; trial registration number: ClinicalTrials.gov, Identifier: NCT02977819). All participants provided signed informed consent prior to participation.

**Table 1 :**
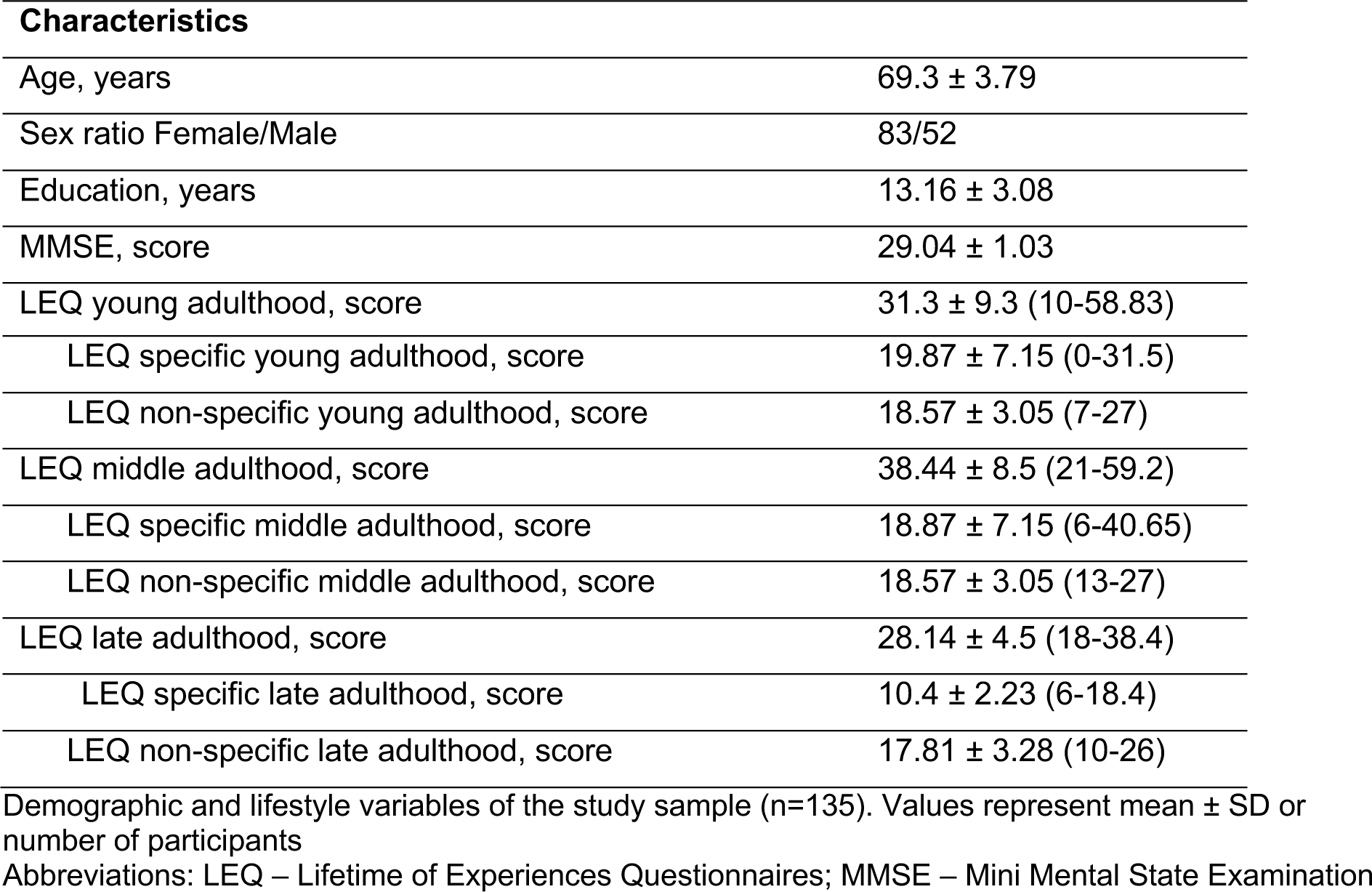
Participants’ characteristics.

| **Characteristics** |  |
| --- | --- |
| Age, years | 69.3 ± 3.79 |
| Sex ratio Female/Male | 83/52 |
| Education, years | 13.16 ± 3.08 |
| MMSE, score | 29.04 ± 1.03 |
| LEQ young adulthood, score | 31.3 ± 9.3 (10-58.83) |
| LEQ specific young adulthood, score | 19.87 ± 7.15 (0-31.5) |
| LEQ non-specific young adulthood, score | 18.57 ± 3.05 (7-27) |
| LEQ middle adulthood, score | 38.44 ± 8.5 (21-59.2) |
| LEQ specific middle adulthood, score | 18.87 ± 7.15 (6-40.65) |
| LEQ non-specific middle adulthood, score | 18.57 ± 3.05 (13-27) |
| LEQ late adulthood, score | 28.14 ± 4.5 (18-38.4) |
| LEQ specific late adulthood, score | 10.4 ± 2.23 (6-18.4) |
| LEQ non-specific late adulthood, score | 17.81 ± 3.28 (10-26) |

Demographic and lifestyle variables of the study sample (n=135). Values represent mean ± SD or number of participants
Abbreviations: LEQ – Lifetime of Experiences Questionnaires; MMSE – Mini Mental State Examination

### Lifetime of Experiences Questionnaire (LEQ)

The Lifetime of Experiences Questionnaire (LEQ) (Valenzuela & Sachdev, 2007) was adapted in French (Ourry et al., 2021) and used to assess lifestyle across the life-course. The LEQ was self-completed by participants. To limit missing data, questionnaires were checked by a neuropsychologist or a psychometrist; when missing values were identified, the information was collected via phone calls. The questionnaire assesses participants’ engagement in various lifestyle activities across three life periods*: young adulthood* (LEQ-young, 13–30 years), *midlife* (LEQ-midlife, 30–65 years), and *late-life* (LEQ-late, 65 years to present date). For each period, both specific and non-specific activities were assessed. More specifically, specific scores of the LEQ represent the educational experience (primary, secondary and post-secondary education) in *young adulthood*, occupation and managerial experience in *midlife*, and engagement in social, leisure and information-seeking behaviours in *late-life*. Non-specific activities were similar for each life period and assessed participation in various leisure activities including visits to family or friends, music practice, participation in artistic activities, physical activities (of mild, moderate or high intensity), reading, second language practice and travelling. Lifestyle engagement in *young adulthood*, *midlife* and *late-life* were calculated by summing specific and non-specific scores. A total score reflecting lifelong lifestyle engagement was obtained by summing the scores of the three life periods (LEQ-total). Higher scores or sub-scores reflect greater engagement in lifestyle activities.

To allow for comparison between periods (LEQ-young *versus* LEQ-midlife *versus* LEQ-late) or activity types (specific *versus* non-specific), the different LEQ sub-scores were standardized into z-scores.

### Neuroimaging Data

#### Data Acquisition

Participants were all scanned on the same MRI (Philips Achieva 3.0T scanner) and PET (Discovery RX VCT 64 PET-CT scanner; General Electric Healthcare) cameras at the Cyceron Center (Caen, France).

High-resolution T1-weighted anatomical volumes using a three-dimensional fast-field echo sequence (Sagittal 3D-FFE, TR/TE = 7.1/3.3ms, FOV = 256×256mm2, 180 slices, 1×1×1mm3) and a three-dimensional fluid-attenuated inversion recovery image (FLAIR; sagittal; TR/TE 4.8/272ms; inversion time = 1.650ms; FOV = 250×250mm2, 180 slices, voxel size: 0.98×0.98×1mm^3^) were acquired.

FDG-PET scans were acquired after a 6-hour fasting period in 92 participants. After a 30-minute resting period in a quiet and dark environment, ∼180MBq of F^18^-FDG were intravenous injected as a bolus. A 10-minute scan was acquired 50 minutes after injection.

A dual-phase Florbetapir-PET scan was performed to measure brain perfusion (early-acquisition) and amyloid burden (late acquisition). More specifically, a first 10-minute scan started at the time of the intravenous injection of ∼4MBq/kg of F^18^-florbetapir (early-Florbetapir-PET), and a second 10-minute scan was acquired 50 minutes after the injection (late-Florbetapir-PET). Florbetapir-PET acquisitions was missing in one participant and early acquisition was missing in one additional participant due to a scanner issue, such that data was available in 133 participants for early-Florbetapir-PET and 134 for late-Florbetapir-PET.

#### Images processing

T1-weighted images were segmented using the Statistical Parametric Mapping (SPM12) software’s multiple channels segmentation procedure (http://www.fil.ion.ucl.ac.uk/spm/software/spm12) with the corresponding FLAIR images. Grey matter (GM) segments were spatially normalized to the Montreal Neurological Institute (MNI) template and modulated to correct for non-linear warping effects. Medial temporal lobe subregions were also segmented based on T1-weighted MRI images in native-space with the *Automatic Segmentation of Hippocampal Subfields* (ASHS) software (ASHS-T1 atlas; (Yushkevich et al., 2014)).

Each PET image was co-registered onto its corresponding MRI and normalized to the MNI template by applying the deformation parameters defined from the T1-weighted normalization procedure (see above). Images were then quantitatively normalized using the cerebellar GM as the reference region, resulting in standardized uptake value ratio (SUVR).

Analyses were conducted both on AD-signature regions of interest (ROI; Figure 1B) and voxel-wise. To investigate AD-specific ROI, hippocampal volume was obtained from the ASHS segmentation. Left and right volumes were averaged and raw bilateral volumes were then normalized by the total intracranial volume (TIV) to account for interindividual variability in head size (volume x100/TIV). Metabolic- and amyloid-signature ROIs were defined as previously (Besson et al., 2015). More specifically, average glucose metabolism and cerebral perfusion was extracted from temporo-parietal regions and neocortical amyloid burden was extracted from the entire gray matter, except the cerebellum, occipital and sensory motor cortices, hippocampi, amygdala and basal nuclei (La Joie et al., 2013).

Normalized MRI and PET images were finally smoothed with a Gaussian kernel of 10mm full width half maximum to allow for voxel-wise analyses.

#### Statistical analyses

We first assessed the associations between lifestyle at each life period and AD-sensitive regions for each imaging modality (Figure 1B). Then, we extended our analyzes to the whole brain and used voxel-wise multiple regressions to assessed the associations between lifestyle at each life period and neuroimaging (Figure 1C1). When an association between a given period and neuroimaging was evidenced, we assessed the specificity of this result. More specifically, we compared voxel-wise, within the cluster(s) previously evidenced (inclusive mask), the association between this life period and neuroimaging to the association with the association for the two other life periods (post-hoc contrast from the multiple regression model; Figure 1C2). The same analyses were replicated without the inclusive mask and results were similar; as a result only results using the inclusive mask will be presented. The average volume/SUVR from these regions was then extracted to conduct the next analyses. A 2-by-2 correlation coefficient comparisons between each life period and extracted volume/SUVR using the *cocor* package (Diedenhofen & Musch, 2015) were conducted (Figure 1C3). Finally, we investigated whether one type of activity (specific *versus* non-specific activities) could drive these effects (Figure 1C4) and, for the significant life period, conducted correlation coefficient comparisons between specific versus non-specific activities and the extracted volume/SUVR using the *cocor* package.

All analyses were conducted controlling for age, sex and the other life periods (or the other sub-score for analyses involving the comparisons between specific and non-specific sub-scores). The *cocor* method doesn’t allow the inclusion of covariates, as a result, models were replicated using residuals from the linear association between brain outcomes and age, sex and other life periods. Results were similar with or without residuals and only results without residuals will be presented. Voxelwise analyses were conducted on SPM12 and other statistical analyses were conducted using R studio. Results were considered significant at p<.005 and k>100 for voxel-wise analyses, and p<.05 for the other analyses.

## Results

### Association between lifestyle at different life periods and AD-sensitive regions

Multiple linear regressions revealed no association between AD-sensitive regions and LEQ scores at any life period (ps>0.25, Table 2).

**Table 2 :** Association between LEQ scores for each life period and AD-sensitive regions.

| AD-sensitive regions | LEQ-young |  |  | LEQ-midlife |  |  | LEQ-late |  |  |
| --- | --- | --- | --- | --- | --- | --- | --- | --- | --- |
|  | b | t | p | b | t | p | b | t | p |
| Hippocampal volume | .004 | .14 | .89 | -.01 | -.50 | .62 | -.007 | -.26 | .80 |
| Temporo-parietal glucose metabolism | .015 | .40 | .69 | .016 | 0.77 | .44 | -.018 | -.38 | .70 |
| Temporo-parietal cerebral perfusion | <.001 | .002 | .99 | <.001 | -0.81 | .42 | <.001 | .38 | .71 |
| Neocortical amyloid burden | .009 | .55 | .58 | -.02 | -1.16 | .25 | -.01 | -.75 | .45 |

Statistical values were obtained from multiple linear regressions including LEQ z-scores for the three life periods (in the same models) as predictors of neuroimaging, controlling for age, sex
Abbreviation: LEQ, Lifetime of Experiences Questionnaire

### Whole-brain association between lifestyle for each life period and neuroimaging

Voxelwise multiple regressions, including LEQ scores for the three life periods in the same models, revealed no association between the LEQ-young score and neuroimaging.

LEQ-midlife score was associated with higher GMv in the left superior anterior cingulate and middle cingulate, middle and inferior temporal gyrus, left middle frontal, inferior frontal gyrus, insula and Heschl’s gyrus as well as the right superior frontal gyrus (Figure 2A). LEQ-midlife was also associated with lower amyloid burden in the right middle cingulate cortex, middle temporal pole and cuneus (Figure 2B). No association was found with LEQ-midlife and glucose metabolism or cerebral perfusion.

**Figure 2:**
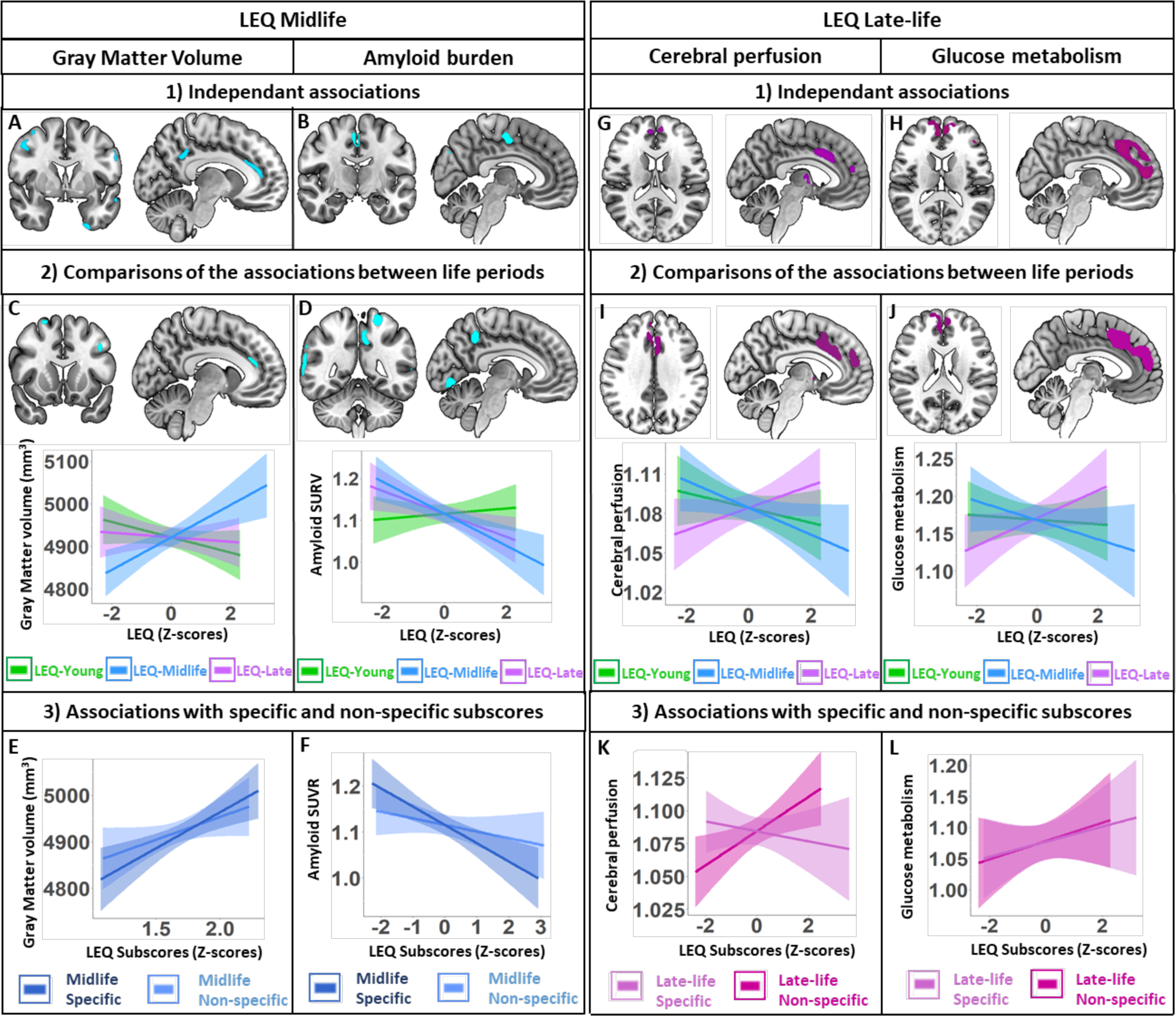
Associations between midlife and late-life LEQ scores and brain integrity. Associations between LEQ-midlife and Gray Matter volume (A) and amyloid burden (B) controlled for age, sex and LEQ-late and LEQ-young scores. Regions showing greater association with LEQ-midlife compared to the other two periods for gray matter volume (C) and amyloid burden (D) and associated graphic representations. Comparison of the association between LEQ-midlife specific (dark blue) *versus* non-specific subscores (light blue) and GMv (E) and amyloid burden (F) within the regions showing a greater association with LEQ-midlife scores. Associations between LEQ-late and Cerebral Perfusion (G) and Glucose metabolism (H) controlled for age, sex and LEQ-midlife and LEQ-young scores. Regions showing greater association with LEQ-late compared to the other two periods for cerebral perfusion (I) and glucose metabolism (J) and associated graphic representations. Comparison of the association between LEQ-late specific (light pink) and non-specific subscores (dark pink) and cerebral perfusion (K) and glucose metabolism (L) within the regions showing a greater association with LEQ-late scores.

Voxelwise multiple regressions showed that the LEQ-late score was associated with higher cerebral perfusion in the left superior anterior cingulate, middle cingulate, the superior medial frontal gyri and (Figure 2G). The LEQ-late score was also associated with higher glucose metabolism in the anterior and middle cingulate, the superior medial frontal gyri and the left thalamus (Figure 2H). No association was found between LEQ-late and GMv or amyloid burden.

### Association between LEQ-midlife and GMv

#### Comparison between LEQ-midlife and the two other periods

To further assess the specificity of the association between LEQ-midlife scores and GMv, we directly assessed whether this association was stronger than the association with the two other life periods. Analyses were restricted to the brain regions found to be associated with LEQ-midlife (Figure 2A; inclusive mask at p<.05 uncorrected). Voxel-wise comparisons revealed that the positive association between LEQ-midlife scores and GMv was stronger than with the two other periods in the left superior anterior cingulate, middle and inferior temporal gyrus, inferior frontal gyrus, insula and Heschl’s gyrus, as well as the right superior frontal gyrus (Figure 2C).

The comparison of correlation coefficients between the average GMv of these regions and LEQ scores further indicates that GMv was more positively correlated with LEQ-midlife than it was to LEQ-young (p<.001) and LEQ-late score (p<.001), while the correlations with LEQ-young and LEQ-late scores did not differ between each other (supplementary table 1A).

#### Comparison between LEQ-midlife specific and non-specific subscores

Multiple linear regression between the average GMv in the same regions and the LEQ-midlife specific and non-specific subscores revealed that specific midlife activities were positively associated with GMv (b=150.2, p=.006) while non-specific activities were not (b=50.9, p=.35, Figure 2E). Comparison of correlation coefficients between LEQ-midlife specific and non-specific subscores indicated that their correlations with GMv did not differ (supplementary table 4).

### Association between LEQ-midlife and amyloid burden

#### Comparison between LEQ-midlife and the two other periods

Analyses restricted to the brain regions previously highlighted (Figure 2B; inclusive mask at p<.05 uncorrected) revealed that the negative association between LEQ-midlife score and amyloid burden was stronger than with the two other periods in the left precuneus and superior parietal gyrus, as well as in the right middle and superior temporal gyrus (Figure 2D).

The comparison of correlation coefficients between the average amyloid burden within these regions and LEQ scores further revealed that amyloid burden was more strongly associated with LEQ-midlife than LEQ-young (p<.001), but did not differ from the correlation with LEQ-late (p=.83) (supplementary table 2B). The correlation between amyloid burden with LEQ-late score was also higher than with LEQ-young (p=.004) (supplementary table 2B).

#### Comparison between LEQ-midlife specific and non-specific subscores

Amyloid burden (in the regions previously highlighted) was negatively associated with LEQ-midlife specific subscore (b=-.13, p<.001), but not with the non-specific subscore (b=.02, p=0.71) (Figure 2F). The direct comparison of correlation coefficients between LEQ-midlife specific and non-specific subscores further indicates that the specific score was more strongly associated with amyloid burden than the non-specific score (p<.05) (supplementary table 4).

### Association between LEQ-late-life and cerebral perfusion

#### Comparison between LEQ-late and the two other periods

Voxel-wise comparisons showed that the positive association previously evidenced between LEQ-late score and cerebral perfusion was stronger than it was with the two other periods in the left superior anterior cingulate, middle cingulate, superior medial frontal gyri and thalamus (Figure 2I).

The comparison of correlation coefficients between the average perfusion within these regions and LEQ scores further indicates that LEQ-late was more positively associated with cerebral perfusion than LEQ-young (p<.05) and LEQ-midlife (p<.001) scores, while the correlation coefficients for LEQ-young and LEQ-midlife did not differ between each other (p=.19, supplementary table 3A).

#### Comparison between LEQ-late specific and non-specific subscores

Multiple linear regressions between LEQ-late subscores and the average cerebral perfusion (in the regions previously highlighted) revealed a positive association between the LEQ-late non-specific subscore and cerebral perfusion (b=0.01, p=.003) but no association with the LEQ-late specific subscore (b=-.002, p=.73) (Figure 2K). Comparison of correlation coefficients further indicated that LEQ-late non-specific subscore was more strongly associated with cerebral perfusion than the specific subscore (p<.05, supplementary table 4A).

### Association between LEQ-late-life and glucose metabolism

#### Comparison between LEQ-late and the two other periods

The positive association found between glucose metabolism with LEQ-late score was stronger than it was with the two other periods in the anterior and middle cingulate and superior medial frontal gyri (Figure 2J).

The comparison of correlation coefficients revealed that the average glucose metabolism within these regions was more strongly associated with LEQ-late than it was with the LEQ-young (p<.05) or LEQ-midlife subscores (p<.001, supplementary table 2B). No difference was found between LEQ-young and LEQ-midlife (p>0.05, supplementary table 2B)

#### Comparison between LEQ-late specific and non-specific subscores

Multiple linear regression between glucose metabolism (extracted from the regions detailed above) and LEQ-late sub-scores showed that only LEQ-late specific subscore was associated with glucose metabolism (b=.16, p=.05) (Figure 2L). Comparison of correlation coefficients showed that the association with glucose metabolism did not differ between LEQ-late specific or non-specific subscores (p=.20, supplementary table 4B).

## DISCUSSION

The aim of this study was to better understand the association between lifestyle across adulthood and brain markers of ageing and AD in cognitively unimpaired older adults. While no associations were found within AD-sensitive regions, lifestyle was differently associated with the integrity of other brain regions according to the life period considered. Interestingly, these other regions such as the anterior cingulate cortex, the frontal areas and the precuneus have been associated with reserve and resilience(Arenaza-Urquijo et al., 2019). Indeed, some studies have highlighted their associations with cognitive resilience (Arenaza-Urquijo et al., 2019) and reserve proxies in healthy (Greenwood, 2000; Pardo et al., 2007, 2020) as well as pathological ageing (Colangeli et al., 2016; Pezzoli et al., 2023; Álvares-Pereira et al., 2022; Menardi et al., 2018).

Our study suggests that lifestyle engagement during midlife is more specifically associated with greater structural integrity, notably in the anterior cingulate cortex, and lower amyloid burden in the precuneus, as compared to the other life periods. These results are consistent with the literature as midlife engagement in physical and cognitive activities have been described as having a protective role on the brain structure in older adults, both structurally (Hofman et al., 2023). A diverse lifestyle at midlife has also been associated with lower amyloid burden and less white matter lesions in healthy older adults (Cadar et al., 2012; Chan et al., 2018; Kim et al., 2018). While longitudinal studies are mandatory to better interpret these findings, they might indicate that midlife lifestyle engagement could promote brain reserve or brain maintenance (Stern et al., 2020, 2023) allowing older adult to maintain volume and resist to amyloid accumulation. Although the findings related to gray matter volume were not related to a specific type of activity, the association with lower amyloid burden was mainly explained by the specific score during midlife, reflecting occupation complexity, especially the managerial aspect. While on a different outcome, this result is in line with a study showing that cognitively unimpaired older adults working with a managerial experiment during midlife showed a smaller reduction in hippocampal volume (Suo et al., 2017). Altogether, these results suggest that engagement in complex mental activities in midlife, especially occupations, could have long-term effect on brain health and promote reserve in old age.

In addition, lifestyle engagement during late-life was associated with greater brain perfusion and glucose metabolism in frontal median regions, including the anterior and middle cingulate cortex and superior medial frontal gyri. This association was stronger than the association with young adulthood and midlife engagements, suggesting that late-life/current lifestyle might be the period at which lifestyle most closely related to brain function. These results align with previous studies showing the advantages, for older adults, of having a healthy and rich lifestyle in late-life on cognition and brain health, including greater physical activity (Matura et al., 2017; Scarmeas et al., 2003; Shah et al., 2014) or cognitive activities (Casaletto et al., 2020; Wu et al., 2023). Notably, cognitive engagement in late adulthood has been associated with higher glucose metabolism (Lyons et al., 2018). Such higher medial frontal perfusion/metabolism could be the reflect of increased cognitive reserve (Stern et al. 2023), more efficient or active networks, promoted by current engagement in complex mental activities. Assessing the relative influence of late-life specific and non-specific activities on these associations, results were rather contrasted indicating a greater influence of late-life non-specific activities for cerebral perfusion and of late-life specific activities for glucose metabolism. Of importance, and contrary to young adulthood and midlife, specific and non-specific activities after retirements are relatively similar. While more fine-grained assessment of activity types (e.g. social, artistic, cognitive, physical engagement) could help understand these results, it appears that both types of activities are associated with greater brain function in late-life.

Surprisingly, no association between lifestyle in young adulthood and brain integrity was found in our study. However, a large body of studies has established an association between young adulthood lifestyle engagement, especially the level of education, and both structural and functional benefits on the brain (Arenaza-Urquijo et al., 2013, 2017; Mintzer et al., 2019). Nevertheless, another study (Nyberg et al., 2021) underlined no relationship between the level of education and brain aging, notably with the rate of change in atrophy-prone cortical regions. While studies are still needed to better understand these inconsistencies, the lack of association in our study might be explained by the fact that participants who participated in our trial were highly-educated, which might reduce our power to detect any effect.

Overall, lifestyle at midlife could have long-term effects on brain health by improving gray matter volume and reduce amyloid burden in brain regions associated with reserve and ageing. Our study suggests that complex mental activities at midlife, particularly occupation, are more strongly associated with structural and molecular markers of brain integrity in older adults and could promote resistance to brain alterations. On the other hand, late-life (*i.e.* current) activities were more strongly associated with brain function, suggesting that current older adults’ lifestyle could help maintain a more active and functional brain.

## STRENGTHS AND LIMITATIONS

One of main strengths of the study is the assessment of lifestyle across adulthood (past and current lifestyle) in a very well characterized sample of older adults. Such data not only allows to evaluate the effect of lifestyle at each life period, but it allows to control analyses for lifestyle engagement at the other period and better disentangle the independent and relative influence of the different life periods. In addition, the multimodal aspect of this study allows to reach a more integrated view of the association between lifestyle and brain health and better touch the underlying mechanism. This study also has limitations. The Lifetime of Experiences Questionnaires (LEQ) is based on a subjective and retrospective evaluation of the participants. This retrospective aspect can lead to uncertainties in the answers. Nevertheless, the LEQ has been validated previously using test-retest (Ourry et al., 2021) and the inclusion of only cognitively unimpaired older adults in the study should limit recollection bias. The transversal design of this study prevents causality and larger longitudinal studies are needed to better understand the mechanisms by which lifestyle could promote reserve across the life-course. Finally, our participants are highly educated and motivated with a relatively healthy lifestyle and future study, with more diverse population, are needed.

## CONCLUSION

Lifestyle at different life periods might have complementary benefits on distinct markers of brain health in late adulthood. Interestingly, no associations were found on AD-sensitive regions but in brain regions relevant to reserve and aging. Lifestyle at midlife might help to promote brain reserve (i.e. gray matter volume) and resistance to brain pathology (i.e. amyloid) in late-life, while current/late-life lifestyle would be more related to brain function. In line with reserve theories, a rich lifestyle throughout life seems to be associated with better brain reserve/maintenance and cognitive reserve, and would enable older adults to maintain brain function as well as better brain integrity. This study provides a better understanding of the factors influencing cerebral aging and may guide recommendations in neurodegenerative disease prevention, in particular by identifying the life period(s) to target.

## Data Availability

The data underlying this report are made available on request following a formal data sharing agreement and approval by the consortium and executive committee (https://silversantestudy.eu/2020/09/25/data-sharing)

## ACKNOWLEDGEMENTS

The authors thank all the Medit-Ageing Research Group members and the Cyceron MRI-PET staff members for their help with recruitment, data acquisition, or administrative support; the EUCLID team, the sponsor (INSERM) and the participants of the study for their contribution. The full Medit-Ageing Research Group is listed in the Appendix.

