## Supplementary Material for "Association between lifestyle at different life periods and brain integrity in older adults"

This document contains:

- The Medit-Ageing research Group list
- Supplementary Figure and tables

### Collaborators - Medit-Ageing Research Group

| Name | Location | Role | Contribution |
|---|---|---|---|
| Allais Florence, BA | EUCLID/F-CRIN Clinical Trials Platform, Bordeaux, France | Data manager | Data management |
| André Claire, PhD | Institut National de la Santé et de la Recherche Médicale, Caen, France | PhD student | Acquisition, analysis, or interpretation of data |
| Arenaza Urquijo Eider, PhD | Institut National de la Santé et de la Recherche Médicale, Caen, France | Postdoctoral researcher | Study design; acquisition, analysis, or interpretation of data |
| Baez Lugo Sebastian, MSc | University of Geneva, Geneva, Switzerland | PhD student | Acquisition, analysis, or interpretation of data |
| Bejanin Alexandre, PhD | Institut National de la Santé et de la Recherche Médicale, Caen, France | Postdoctoral researcher | Acquisition, analysis, or interpretation of data |
| Botton Maelle, MSc | Institut National de la Santé et de la Recherche Médicale, Caen, France | Neuropsychologist | Acquisition, analysis, or interpretation of data |
| Champetier Pierre, PhD | Institut National de la Santé et de la Recherche Médicale, Caen, France | PhD student | Acquisition, analysis, or interpretation of data |
| Chauveau Léa, MSc | Institut National de la Santé et de la Recherche Médicale, Caen, France | PhD student | Acquisition, analysis, or interpretation of data |
| Chételat Gaël, PhD | Institut National de la Santé et de la Recherche Médicale, Caen, France | Coordinator,Work Package Leader | Obtained funding, study design |
| Chocat Anne, MD | Institut National de la Santé et de la Recherche Médicale, Caen, France | Neurologist | Investigating doctor |
| Collette Fabienne, PhD | University of Liege, Liege, Belgium | Group leader | Obtained funding, study design |
| Dautricourt Sophie, MD,PhD | Institut National de la Santé et de la Recherche Médicale, Caen, France | PhD student | Acquisition, analysis, or interpretation of data |
| de Flores Robin, PhD | Institut National de la Santé et de la Recherche Médicale, Caen, France | Postdoctoral researcher | Acquisition, analysis, or interpretation of data; administrative, technical, or material support |
| de la Sayette Vincent, MD, PhD | Centre Hospitalier Universitaire de Caen, Caen, France | Neurologist | Principal Investigating doctor |
| Delarue Marion, MSc | Institut National de la Santé et de la Recherche Médicale, Caen, France | Neuropsychologist | Acquisition, analysis, or interpretation of data |
| Demnitz-King Harriet, MSc | University College London, United Kingdom | PhD student | Acquisition, analysis, or interpretation of data |
| Egret Stéphanie, MSc | Institut National de la Santé et de la Recherche Médicale, Caen, France | Neuropsychologist | Acquisition, analysis, or interpretation of data |
| El Sadawy Rawda, MSc | Institut National de la Santé et de la Recherche Médicale, Caen, France | Neuropsychologist | Acquisition, analysis, or interpretation of data |

| | | | |
|---|---|---|---|
| Espérou Hélène, MD | Institut National de la Santé et de la Recherche Médicale, Paris, France | Group leader | Sponsor |
| Fauvel Séverine, BA | Institut National de la Santé et de la Recherche Médicale, Caen, France | Technician | Acquisition, analysis, or interpretation of data |
| Felisatti Francesca, MSc | Institut National de la Santé et de la Recherche Médicale, Caen, France | PhD student | Acquisition, analysis, or interpretation of data |
| Ferment Victor, MSc | Institut National de la Santé et de la Recherche Médicale, Caen, France | Technician | Acquisition, analysis, or interpretation of data |
| Ferrand Devouge Eglantine, MD, MSc | Institut National de la Santé et de la Recherche Médicale, Caen, France | Neurologist | Investigating doctor |
| Frison Eric, MD, PhD | EUCLID/F-CRIN Clinical Trials, Platform, Bordeaux, France | Methodologist | Acquisition, analysis, or interpretation of data |
| Gonneaud Julie, PhD | Institut National de la Santé et de la Recherche Médicale, Caen, France | Group leader | Obtained funding, study design |
| Hamel Anaïs, MSc | Institut National de la Santé et de la Recherche Médicale, Caen, France | PhD student | Acquisition, analysis, or interpretation of data |
| Haudry Sacha, MSc | Institut National de la Santé et de la Recherche Médicale, Caen, France | PhD student | Acquisition, analysis, or interpretation of data |
| Hébert Oriane, MSc | Institut National de la Santé et de la Recherche Médicale, Caen, France | Neuropsychologist | Acquisition, analysis, or interpretation of data |
| Heidmann Marc, MSc | Institut National de la Santé et de la Recherche Médicale, Lyon, France | PhD student | Acquisition, analysis, or interpretation of data |
| Kuhn Elizabeth, PhD | Institut National de la Santé et de la Recherche Médicale, Caen, France | PhD student | Acquisition, analysis, or interpretation of data |
| Klimecki Olga, PhD | University of Geneva, Geneva, Switzerland | Group leader | Obtained funding; study design |
| Landeau Brigitte, MSc | Institut National de la Santé et de la Recherche Médicale, Caen, France | Neuroimaging developer (engineer) | Acquisition, analysis, or interpretation of data |
| Ledu Gwendoline, MSc | Institut National de la Santé et de la Recherche Médicale, Caen, France | Clinical research assistant | Acquisition, analysis, or interpretation of data; administrative, technical, or material support |
| Lefranc Valérie, BA | Institut National de la Santé et de la Recherche Médicale, Caen, France | Clinical research assistant | Acquisition, analysis, or interpretation of data; Administrative, technical, or material support |
| Lutz Antoine, PhD | Institut National de la Santé et de la Recherche Médicale, Lyon, France | Group leader | Obtained funding; study design |
| Marchant Natalie, PhD | University College London, United Kingdom | Group leader | Obtained funding; study design |
| Mezenge Florence, BA | Institut National de la Santé et de la Recherche Médicale, Caen, France | Neuroimaging engineer assistant | Acquisition, analysis, or interpretation of data |

| | | | |
|---|---|---|---|
| Moulinet Inès, PhD | Institut National de la Santé et de la Recherche Médicale, Caen, France | PhD student | Acquisition, analysis, or interpretation of data |
| Ourry Valentin, PhD | Institut National de la Santé et de la Recherche Médicale, Caen, France | PhD student | Acquisition, analysis, or interpretation of data |
| Palix Cassandre, MSc | Institut National de la Santé et de la Recherche Médicale, Caen, France | PhD student | Acquisition, analysis, or interpretation of data |
| Paly Léo, MSc | Institut National de la Santé et de la Recherche Médicale, Caen, France | Neuropsychologist | Acquisition, analysis, or interpretation of data |
| Poisnel Géraldine, PhD | Institut National de la Santé et de la Recherche Médicale, Caen, France | Group leader | Obtained funding; study design |
| Quillard Anne, MD | Institut National de la Santé et de la Recherche Médicale, Caen, France | Neurologist | Investigating doctor |
| Rauchs Géraldine, PhD | Institut National de la Santé et de la Recherche Médicale, Caen, France | Researcher | Study design |
| Rehel Stéphane, PhD | Institut National de la Santé et de la Recherche Médicale, Caen, France | PhD student | Acquisition, analysis, or interpretation of data |
| Requier Florence, MSc | University of Liege, Liege, Belgium | PhD student | Acquisition, analysis, or interpretation of data |
| Salmon Eric, MD,PhD | University of Liege, Liege, Belgium | Group leader | Obtained funding; study design |
| Smith Rhonda, MSc | Minerva Health & Care Communications Ltd, Andover, United Kingdom | Group leader | Communication, dissemination |
| Tomadesso Clémence, PhD | Institut National de la Santé et de la Recherche Médicale, Caen, France | PhD student | Acquisition, analysis, or interpretation of data |
| Touron Edelweiss, PhD | Institut National de la Santé et de la Recherche Médicale, Caen, France | PhD student | Acquisition, analysis, or interpretation of data |
| Turpin Anne-Laure, MSc | Institut National de la Santé et de la Recherche Médicale, Caen, France | PhD student | Acquisition, analysis, or interpretation of data |
| Vuilleumier Patrik, MD | University of Geneva, Geneva, Switzerland | Group leader | Obtained funding; study design |
| Whitfield Tim, PhD | University College London, United Kingdom | PhD student | Acquisition, analysis, or interpretation of data |
| Wirth Miranka, PhD | Deutsches Zentrum für Neurodegenerative Erkrankungen, Dresden, Germany | Group leader | Study design |

**Supplementary Figure 1:** Flow chart

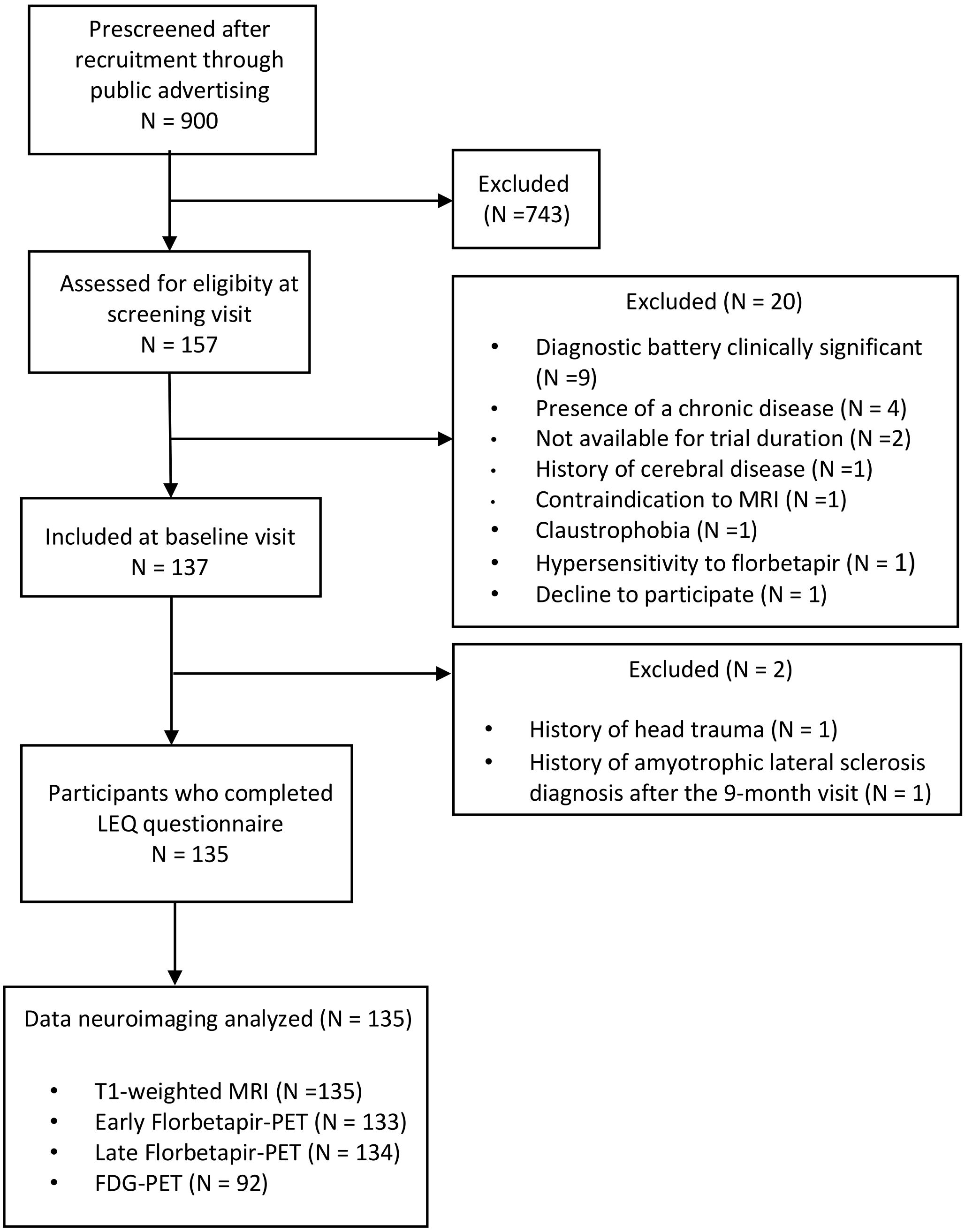

**Supplementary Table 1**: Correlation coefficient comparisons of the association between lifestyle at each life periods and (a) gray matter volume and (b) amyloid burden

**a) Gray Matter volume (GMv)**

| | **GMv ~ LEQ-Midlife** vs **GMv ~ LEQ-Late** | **GMv ~ LEQ-Midlife** vs **GMv ~ LEQ-Young** | **GMv ~ LEQ-Late** vs **GMv ~ LEQ-Young** |
|---|---|---|---|
| Pearson and Filon's z (1898) | z = 3.7834, **p = 0.0001** | z = 5.0082, **p = 0.0000** | z = 0.9298, p = 0.1762 |
| Hotelling's t(1940) | t = 3.8162, df = 132, **p = 0.0001** | t = 5.1244, df = 132, **p = 0.0000** | t = 0.9204, df = 132, p = 0.1795 |
| William's t (1959) | t = 3.8134, df = 132, **p= 0.0001** | t = 5.1229, df = 132, **p = 0.0000** | t = 0.9198, df = 132, p = 0.1797 |
| Olkin's z (1967) | z = 3.7834, **p = 0.0001** | z = 5.0082, **p = 0.0000** | z = 0.9298, p = 0.1762 |
| Dunn and Clark's z (1969) | z = 3.6868, **p = 0.0001** | z = 4.8192, **p = 0.0000** | z = 0.9298, p = 0.1762 |
| Stanley and Hills' (1970) | t = 3.8162, df = 132, **p = 0.0001** | t = 5.1243, df = 132, **p = 0.0000** | t = 0.9204, df = 132, p = 0.1795 |
| Meng, Rosenthal, and Rubin's z (1992) | z = 3.6181, **p = 0.0001** | z = 4.6734, **p = 0.0000** | z = 0.9168, p = 0.1796 |
| Hittner, May, Silver's (2003) | z = 3.6517, **p = 0.0001** | z = 4.7478, **p = 0.0000** | z = 0.9175, p = 0.1794 |
| Zou's (2007) confidence interval | 0.1492 0.4808 | 0.2448 0.5682 | -0.1048 0.2876 |

**b) Amyloid burden**

| | **Amyloid burden ~ LEQ-Midlife** vs **Amyloid burden ~ LEQ-Late** | **Amyloid burden ~ LEQ-Midlife** vs **Amyloid burden ~ LEQ-Young** | **Amyloid burden ~ LEQ-Late** vs **Amyloid burden ~ LEQ-Young** |
|---|---|---|---|
| Pearson and Filon's z (1898) | z = -0.9725, p = 0.8346 | z = -4.0523, **p = 0.0000** | z = -2.6338, **p = 0.0042** |
| Hotelling's t(1940) | t = -0.9662, df = 131, p = 0.8321 | t = -4.1019, df = 131, **p = 0.0000** | t = -2.6147, df = 131, **p = 0.005** |
| William's t (1959) | t = -0.9633, df = 131, p = 0.8314 | t = -4.0988, df = 131, **p = 0.0000** | t = -2.6132, df = 131, **p = 0.005** |
| Olkin's z (1967) | z = -0.9725, p= 0.8346 | z = -4.0523, **p = 0.0000** | z = -2.6338, **p = 0.0042** |
| Dunn and Clark's z (1969) | z = -0.9612, p = 0.8318 | z = -3.9408, **p = 0.0000** | z = -2.5765, **p = 0.005** |
| Stanley and Hills' (1970) | t = -0.9662, df = 131, p = 0.8321 | t = -4.1019, df = 131, **p = 0.0000** | t = -2.6147, df = 131, **p = 0.005** |
| Meng, Rosenthal, and Rubin's z (1992) | z = -0.9599, p= 0.8315 | z = -3.8574, **p = 0.0001** | z = -2.5483, **p= 0.0054** |
| Hittner, May, Silver's (2003) | z = -0.9605, p = 0.8316 | z = -3.8983, **p = 0.0000** | z = -2.5622, **p = 0.0052** |
| Zou's (2007) confidence interval | -0.2474 0.0847 | -0.5011 -0.1709 | -0.4469 -0.0616 |

Comparisons of coefficient correlations between life periods for a) GMv and b) amyloid burden were performed on R-Studio. Statistical values were obtained after testing the alternative hypothesis « greater » for GMv and « less » for amyloid burden and were significant when $p<0.05$.

Abbreviations: LEQ : Lifetime of Experiences Questionnaire ; GMv : Gray Matter volume

**Supplementary Table 2**: Correlation coefficient comparisons of the association between lifestyle at each life periods and (a) cerebral perfusion and (b) glucose metabolism

**a) Cerebral perfusion**

| | **Cerebral perfusion ~ LEQ-Late** vs **Cerebral perfusion ~ LEQ-Midlife** | **Cerebral perfusion ~ LEQ-Late** vs **Cerebral perfusion ~ LEQ-Young** | **Cerebral perfusion ~ LEQ-Young** vs **Cerebral perfusion ~ LEQ-Midlife** |
|---|---|---|---|
| Pearson and Filon's z (1898) | z = 3.5665, **p = 0.0002** | z = 2.3046, **p= 0.0106** | z = 0.8734, p-value = 0.1912 |
| Hotelling's t(1940) | t = 3.5800, df = 130, **p = 0.0002** | t = 2.2836, df = 130, **p = 0.012** | t = 0.8650, df = 130, p-value = 0.1943 |
| William's t (1959) | t = 3.5800, df = 130, **p= 0.0002** | t = 2.2835, df = 130, **p = 0.012** | t = 0.8644, df = 130, p-value = 0.1945 |
| Olkin's z (1967) | z = 3.5665, **p= 0.0002** | z = 2.3046, **p= 0.0106** | z = 0.8734, p-value = 0.1912 |
| Dunn and Clark's z (1969) | z = 3.4712, **p= 0.0003** | z = 2.2586, **p = 0.012** | z = 0.8628, p-value = 0.1941 |
| Stanley and Hills' (1970) | t = 3.5800, df = 130, **p= 0.0002** | t = 2.2836, df = 130, **p = 0.012** | t = 0.8650, df = 130, p-value = 0.1943 |
| Meng, Rosenthal, and Rubin's z (1992) | z = 3.4128, **p= 0.0003** | z = 2.2392, **p= 0.0126** | z = 0.8619, p-value = 0.1944 |
| Hittner, May, Silver's (2003) | z = 3.4425, **p = 0.0003** | z = 2.2489, **p= 0.0123** | z = 0.8623, p-value = 0.1943 |
| Zou's (2007) confidence interval | 0.1327 0.4688 | 0.0301 0.4205 | -0.0960 0.2462 |

**b) Glucose metabolism**

| | **Glucose metabolism ~ LEQ-Late** vs **Glucose metabolism ~ LEQ-Midlife** | **Glucose metabolism ~ LEQ-Late** vs **Glucose metabolism ~ LEQ-Young** | **Glucose metabolism ~ LEQ-Midlife** vs **Glucose metabolism ~ LEQ-Young** |
|---|---|---|---|
| Pearson and Filon's z (1898) | z = 3.2850, **p= 0.0005** | z = 1.9051, **p= 0.0284** | z = 1.0815, p-value = 0.1397 |
| Hotelling's t(1940) | t = 3.2914, df = 89, **p= 0.0007** | t = 1.8796, df = 89, **p= 0.0317** | t = 1.0664, df = 89, p-value = 0.1446 |
| William's t (1959) | t = 3.2913, df = 89, **p= 0.0007** | t = 1.8786, df = 89, **p= 0.0318** | t = 1.0661, df = 89, p-value = 0.1446 |
| Olkin's z (1967) | z = 3.2850, **p= 0.0005** | z = 1.9051, **p= 0.0284** | z = 1.0815, p-value = 0.1397 |
| Dunn and Clark's z (1969) | z = 3.1695, **p= 0.0008** | z = 1.8584, **p= 0.0316** | z = 1.0617, p-value = 0.1442 |
| Stanley and Hills' (1970) | t = 3.2913, df = 89, **p= 0.0007** | t = 1.8796, df = 89, **p= 0.0317** | t = 1.0664, df = 89, p-value = 0.1446 |
| Meng, Rosenthal, and Rubin's z (1992) | z = 3.1047, **p= 0.0010** | z = 1.8427, **p= 0.0327** | z = 1.0592, p-value = 0.1447 |
| Hittner, May, Silver's (2003) | z = 3.1378, **p= 0.0009** | z = 1.8505, **p= 0.0321** | z = 1.0605, p-value = 0.1445 |
| Zou's (2007) confidence interval | 0.1287 0.5335 | -0.0125 0.4551 | -0.0934 0.3121 |

Comparisons of coefficient correlations between life periods for a) cerebral perfusion and b) glucose metabolism were performed on R-studio. Statistical values were obtained and significant when p<0.05 after testing the alternative hypothesis « greater » for both functional modalities.

Abbreviations: LEQ : Lifetime of Experiences Questionnaire

**<u>Supplementary Table 3</u>**: Correlation coefficient comparisons of the association between LEQ-midlife specific and non-specific scores and (a) gray matter volume and (b) amyloid burden

| | **a) Gray Matter volume (GMv)**<br>**GMv ~ LEQ-Midlife specific**<br>vs<br>**GMv ~ LEQ-Midlife non-specific** | **b) Amyloid burden**<br>**Amyloid burden ~ LEQ-Midlife specific**<br>vs<br>**Amyloid burden~ LEQ-Midlife non-specific** |
|---|---|---|
| Pearson and Filon's z (1898) | z = 1.0210, p= 0.1536 | z = -2.0977, p= 0.018 |
| Hotelling's t(1940) | t = 1.0154, df = 132, p= 0.1559 | t = -2.0898, df = 131, p= 0.0193 |
| William's t (1959) | t = 1.0104, df = 132, p= 0.1571 | t = -2.0809, df = 131, p= 0.0197 |
| Olkin's z (1967) | z = 1.0210, p= 0.1536 | z = -2.0977, p= 0.018 |
| Dunn and Clark's z (1969) | z = 1.0083, p= 0.1566 | z = -2.0635, p= 0.0195 |
| Stanley and Hills' (1970) | t = 1.0154, df = 132, p= 0.1559 | t = -2.0898, df = 131, p= 0.0193 |
| Meng, Rosenthal, and Rubin's z (1992) | z = 1.0066, p= 0.1571 | z = -2.0490, p= 0.02 |
| Hittner, May, Silver's (2003) | z = 1.0074, p= 0.1569 | z = -2.0556, p= 0.0199 |
| Zou's (2007) confidence interval | -0.0962 0.2985 | -0.3985 -0.0102 |

Comparisons of coefficient correlations between specific and non-specific middle adulthood activities for a) GMv and b) amyloid burden were performed on R-studio. Statistical values were obtained after testing the alternative hypothesis « greater » for GMv and « less » for amyloid burden and were significant when $p<0.05$.

Abbreviations: LEQ : Lifetime of Experiences Questionnaire ; GMv : Gray Matter volume

**<u>Supplementary Table 4</u>**: Correlation coefficient comparisons of the association between LEQ-late-life specific and non-specific scores and (a) cerebral perfusion and (b) glucose metabolism

| | **a) Cerebral perfusion** | **b) Glucose metabolism** |
|---|---|---|
| | **Cerebral perfusion ~ LEQ-LL non-specific**<br>vs<br>**Cerebral perfusion ~ LEQ-LL specific** | **Glucose metabolism ~ LEQ-LL specific**<br>vs<br>**Glucose metabolism ~ LEQ-LL non-specific** |
| Pearson and Filon's z (1898) | z = 2.4369, p= 0.0074 | z = 0.8534, p= 0.1967 |
| Hotelling's t(1940) | t = 2.4212, df = 130, p= 0.0084 | t = 0.8429, df = 89, p= 0.2008 |
| William's t (1959) | t = 2.4195, df = 130, p= 0.0085 | t = 0.8403, df = 89, p= 0.2015 |
| Olkin's z (1967) | z = 2.4369, p= 0.0074 | z = 0.8534, p= 0.1967 |
| Dunn and Clark's z (1969) | z = 2.3890, p= 0.0084 | z = 0.8384, p= 0.2009 |
| Stanley and Hills' (1970) | t = 2.4212, df = 130, p= 0.0084 | t = 0.8429, df = 89, p= 0.2008 |
| Meng, Rosenthal, and Rubin's z (1992) | z = 2.3670, p= 0.009 | z = 0.8370, p= 0.2013 |
| Hittner, May, Silver's (2003) | z = 2.3778, p= 0.0087 | z = 0.8377, p= 0.2011 |
| Zou's (2007) confidence interval | 0.0415 0.4165 | -0.1289 0.3197 |

Comparisons of coefficient correlations between specific and non-specific late adulthood activities for a) cerebral perfusion and b) glucose metabolism were performed on R-studio. Statistical values were obtained after testing the alternative hypothesis « greater » for both functional modalities were significant when $p<0.05$.
